# Single-cell RNA sequencing data reveals rewiring of transcriptional relationships in Alzheimer’s Disease associated with risk variants

**DOI:** 10.1101/2023.05.15.23289992

**Authors:** Gerard A. Bouland, Kevin I. Marinus, Ronald E. van Kesteren, August B. Smit, Ahmed Mahfouz, Marcel J.T. Reinders

**Author notes:** Corresponding authors: Ahmed Mahfouz and Marcel J.T. Reinders.

## Abstract

Understanding how genetic risk variants contribute to Alzheimer’s Disease etiology remains a challenge. Single-cell RNA sequencing (scRNAseq) allows for the investigation of cell type specific effects of genomic risk loci on gene expression. Using seven scRNAseq datasets totalling >1.3 million cells, we investigated differential correlation of genes between healthy individuals and individuals diagnosed with Alzheimer’s Disease. Using the number of differential correlations of a gene to estimate its involvement and potential impact, we present a prioritization scheme for identifying probable causal genes near genomic risk loci. Besides prioritizing genes, our approach pin-points specific cell types and provides insight into the rewiring of gene-gene relationships associated with Alzheimer’s.

## INTRODUCTION

Alzheimer’s Disease (AD) is a progressive neurodegenerative disease characterized by loss of cognitive functions and autonomy, eventually leading to death^1^. Many hypotheses about the etiology of AD exists, e.g. the amyloid-beta (Aβ) cascade hypothesis, the tau hypothesis, the inflammation hypothesis, the oxidative stress hypothesis and more^2,3^, highlighting the complexity of AD. Genome-wide association studies (GWASs) have provided a compendium of genomic loci that are associated with the risk for AD^4–7^. However, understanding how these risk variants contribute to AD etiology remains a challenge. As the number of GWASs is still rising steadily and are increasingly becoming larger in sample size, new genomic risk loci are regularly identified, while studies that generate mechanistic understanding lag behind^8^. Methods such as mendelian randomization^9^ and colocalization^10^ provide insight in causality but fail to provide insight in downstream molecular consequences. Single-cell genomics has made it possible to investigate genetic regulation in distinct cell types and paves the way to new approaches that will provide a more detailed understanding of cell type specific dysregulation in AD, genetics and downstream consequences.

Additionally, scRNAseq has provided insight into cellular heterogeneity and is increasingly used to understand transcriptional differences at a single-cell level^11,12,13^. For AD, several scRNAseq studies have been performed^14,15,16,17,18^ that have generated new insights into AD pathophysiology. Many scRNAseq studies focus on cell type abundance^18^, cell type specific differential expression^14,18^, identifying novel cell types^16^ or exploring cellular differentiation trajectories^19^ – where each cell is kept as an independent entity, while being categorized into distinct cell types. An alternative approach utilizing scRNAseq data involves aggregating multiple measurements of genes within pre-defined cell populations, often delineated by cell type and individual, generating pseudo bulk datasets^20,21^. This approach has successfully been used to identify differential cellular states across conditions^21^, exploring cell type specific responses^22^, and identifying cell type specific gene regulation under genetic control^23^.

While scRNAseq data are well suited for e.g. differential expression analysis (DEA), determining expression correlations in scRNAseq data remains challenging^24^. scRNAseq data is characterized by large numbers of zero counts; the lower the expression, the more abundant the zeros^25^. Consequently, low-expressed genes can appear highly correlated due to just a few paired measurements, while the remaining measurements are zero^26,27^. The pseudo bulk approach provides a solution, as each gene would be represented by the aggregated value within a cell population, delineated by cell type and individual. As such, even low-expressed genes are represented by a robust aggregated value and the correlation is determined by the collinearity between genes across individuals instead of single cells. However, even though most scRNAseq datasets contain large numbers of cells, these are often derived from a small number of individuals, making it challenging to identify meaningful correlations in pseudo bulk data.

To overcome these challenges, we here combined seven previously published AD scRNAseq datasets^14,15,16,17,18^ and generated seven cell type specific pseudo bulk datasets (excitatory neurons, inhibitory neurons, astrocytes, oligodendrocytes, oligodendrocyte progenitor cells (OPCs), microglia and endothelial cells), ranging from 132 to 192 individuals. We used this data to investigate differential correlation^28,29^ of genes between healthy individuals (control, CT) and patients diagnosed with AD. In contrast to DEA, differential correlation analysis (DCA) provides insight in whether transcriptional changes are independent or coordinated and provides insight into dynamic associations of key regulators subject to AD. For each cell type we explored gene-gene correlations that are significantly different in AD compared to CT. Using a network representation of differential correlations, we identified distinct sets of regulatory hubs for each cell type. Using the number of differential correlations to rank genes located near AD genetic risk variants, we prioritized known causal genes and identified potential novel ones. In addition, this approach revealed altered states of biological processes in AD associated with the prioritized genes. Finally, taking advantage of the characteristics of pseudo bulk data, we performed co-expression analysis between genes expressed in excitatory neurons and four other cell types (inhibitory neurons, astrocytes, oligodendrocytes and microglia) to identify pairs of co-expressed genes that are expressed in different cell types.

## RESULTS

### Analysis workflow

The analysis workflow consists of six major components. The first component describes the demographics of the cell type specific pseudo bulk datasets that were composed of seven separate AD scRNAseq datasets (**Fig. 1a**). In the second component, a general overview of differential correlation results between CT and AD is presented (**Fig. 1b**). Then we continue to investigate hubs; genes that have the majority of differential correlations with other genes within each respective cell type **(Fig. 1c)**. In the fourth component, we compare hubs between cell types; are they cell type spcific of shared? Here we test whether shared hubs also share neigbourhoods. In the next component, we use the number of differential correlations of genes genomically located near AD risk variants in a prioritization scheme for identifying putative causal genes and cell types (**Fig. 1d**). In the sixth and final analysis, we perform a co-expression analysis in healthy individuals between genes expressed in excitatory neurons and inhibitory neurons, excitatory neurons and astrocytes, excitatory neurons and oligodendrocytes and finally excitatory neurons and microglia (**Fig. 1e**), thus asking the question whether there are gene-pairs co-expressed across different cell types.

**Figure 1.**
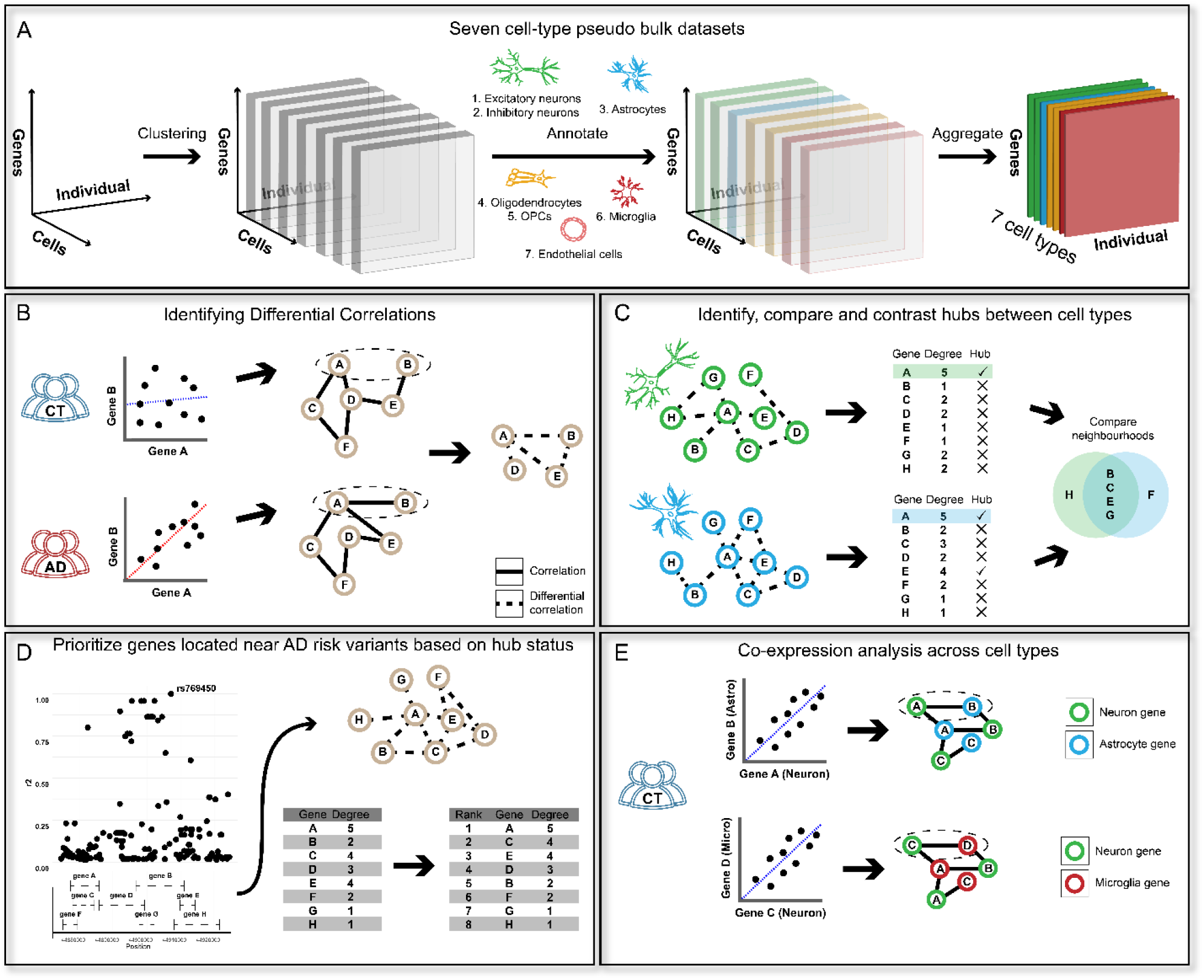
Schematic overview of the analysis workflow. **A** Overview of how the seven cell type specific pseudo bulk datasets were combined. Starting with three dimensions (genes, cells and individuals), the data was clustered along the cells axis. Next, the clusters were annotated, after which the annotated data was aggregated on cell type and individual. This resulted in seven cell type specific datasets with genes defining the rows and the individuals defining the columns. **B** Schematic overview of differential correlations and differential correlation network (DCN). The degree of correlation is first calculated between pairs of genes in both healthy controls (CT) and Alzheimer’s Disease (AD) patients separately, resulting in different co-expression networks. The differential correlation network is defined by the difference between both co-expression networks. **C** Schematic representation of comparing DCNs and the corresponding hubs between cell types. Different cell types have different DCNs and these similarities and differences are identified. Additionally, the network neighborhood genes of the shared hubs is compared. **D** Schematic representation of the gene prioritization scheme. Genes located near AD risk variants are ranked based on their hub status (i.e., based on the number of genes that have an altered association in AD compared to CT). **E** Schematic overview of differential correlation between genes expressed in neurons and genes expressed in astrocytes.

### Demographics of cell type specific datasets

We collected seven scRNAseq Alzheimer’s Disease (AD) datasets, comprised of 1,341,953 cells, and generated seven cell type specific pseudo bulk datasets (*See methods: Aggregation, integration and batch correction*). The excitatory and inhibitory neuron datasets comprise of five datasets (**Table 1**) and consist of 180 individuals, of which 81 were diagnosed with AD and 84 had no cognitive impairment (CT). A total of 15 individuals with mild cognitive impairment (MCI) and/or having other causes for MCI were characterized as other (O) and were excluded from any analyses. The astrocyte, oligodendrocyte and oligodendrocyte progenitor cell (OPC) datasets comprise six datasets and consist of 180 individuals (N_AD_ = 87, N_ct_ = 90, N_O_ = 15). The microglia dataset comprises five datasets and consists of 168 individuals (N_AD_ = 73, N_ct_ = 73, N_O_ = 22). The endothelial cell dataset comprises four datasets and consists of 132 individuals (N_AD_ = 60, N_ct_ = 70, N_O_ = 2).

**Table 1.** Dataset characteristics and demographics

| Cell type | BA9 | BA10 | BA9-46 | LAU | ENT | SEA-AD | BA9-46-Micro | Original No. cells | No. genes | No. subjects | %female | AD | CT | O |
| --- | --- | --- | --- | --- | --- | --- | --- | --- | --- | --- | --- | --- | --- | --- |
| Excitatory Neurons | X | X | X | X |  | X |  | 663,175 | 9,501 | 180 | 51% | 81 | 84 | 15 |
| Inhibitory neurons | X | X | X | X |  | X |  | 268,060 | 3,556 | 180 | 51% | 81 | 84 | 15 |
| Astrocytes | X | X | X | X | X | X |  | 109,713 | 2,615 | 192 | 50% | 87 | 90 | 15 |
| Oligodendrocytes | X | X | X | X | X | X |  | 185,175 | 2,018 | 192 | 50% | 87 | 90 | 15 |
| OPCs | X | X | X | X | X | X |  | 41,611 | 243 | 192 | 50% | 87 | 90 | 15 |
| Microglia |  | X | X | X |  | X | X | 58,443 | 1,356 | 168 | 53% | 73 | 73 | 22 |
| Endothelial cells |  | X | X | X |  | X |  | 15,776 | 368 | 132 | 52% | 60 | 70 | 2 |

### Alzheimer’s Disease is characterized by altered correlations between gene pairs across cell types

To identify altered gene-gene relationships between CT and AD individuals, we performed differential correlation analysis within each cell type. Across all cell types, a total of 374,243 pairs of genes (∼0.65% of all tested pairs, **Fig. 2a**) had altered transcriptional relationships in AD (P_adj_ ≤ 0.01, |Δr| ≥ 0.5). For 253,135 pairs, an increase in correlation coefficient (Δr ≥ 0.5) was observed in AD and for 121,108 pairs a decrease (Δr ≤ -0.5, **Fig.2b**). Most altered relationships were identified in excitatory neurons (n = 313,756, 0.70%), followed by inhibitory neurons (n = 44,974, 0.72%), astrocytes (n = 7,669, 0.22%), microglia (n = 4,061, 0.44%), oligodendrocytes (n = 3,219, 0.61%), endothelial cells (n = 515, 0.77%, **Fig. 2c**) and OPCs (n = 49, 0.17%). We next identified genes that are differentially correlated with age or Braak stage in AD individuals compared to CT (e.g., no correlation between gene expression and age in CT but a positive correlation in AD). Across all cell types, 169 genes were significantly differentially correlated with age (**Fig. 2d-j**, e.g. upregulated with age in CT while downregulated with age in AD) and 215 genes were significantly differentially correlated with Braak stage (**SFig. 1**). *PTPN3* (**Fig. 2k**, r_CT_ = -0.29, r_AD_ = 0.44, P_adj_ = 1.35 × 10^−4^) showed the most extreme changes in association with age in excitatory neurons from AD patients.

**Figure 2.**
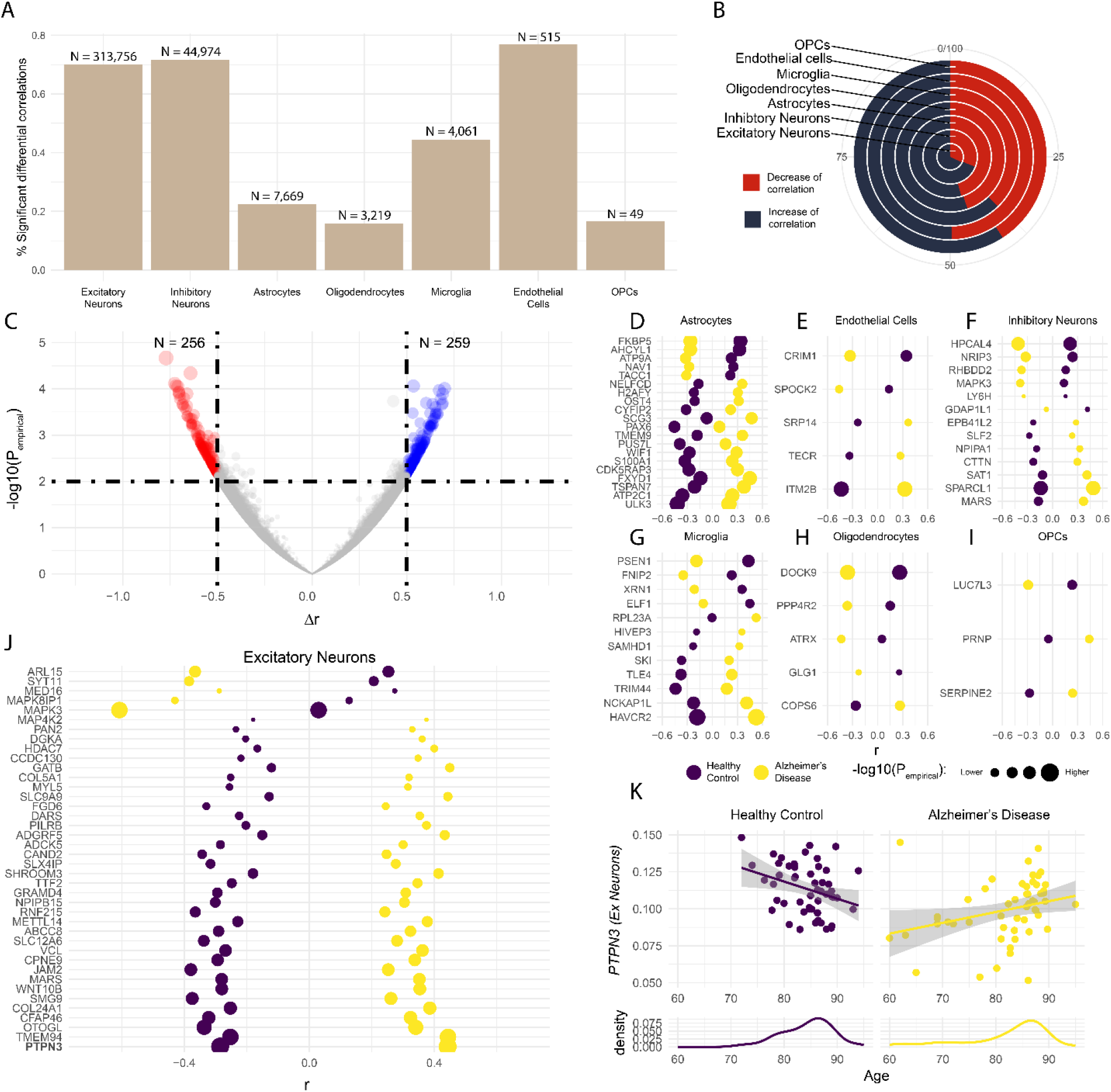
**A** Percentage and number of significant differential correlations for each cell type. **B** For each cell type the percentage of pairs of genes that have an increased or decreased correlation. Increasing meaning a higher correlation coefficient between a pair of genes in individuals with Alzheimer’s Disease (AD) compared to healthy controls (CT), and vice versa. **C** Volcano plot of the differential correlations in endothelial cells. Each dot represents a gene-pair, the x-axis represents the difference in correlation coefficient for the respective pair and the y-axis represents the –log10 empirical P-value. Blue dots are significant differential correlations where an increased correlation coefficient was found in AD. Conversely, red dots are significant differential correlations where a decreased correlation coefficient was found in AD. **D, E, F, G, H, I, J** Plots of the genes that are differentially correlated with age within the respective cell types. The x-axes represent the Spearman’s rank correlation coefficient. Yellow dots represent the correlation coefficient in AD for the respective gene with age and purple dots represent the correlation coefficient in CT for the respective gene with age. The size of the dots correspond to the –log10 p-value. **K** Dot plot for *PTPN3* (excitatory neurons). Each dot is an individual, the x-axes represent the age of the individuals in years and the y-axes the expression of the respective genes in the respective cell types. Yellow dots are individuals with Alzheimer’s and purple dots are healthy controls. Beneath the dot plot a density plot of age is shown.

Next, we investigated whether the observed differential correlations are explained by the differential expression of the genes between CT and AD individuals. Most genes that were differentially correlated were not significantly differentially expressed. Of all genes that were differentially correlated with at least one other gene (n = 18,321), 3,187(∼17%) genes were also significantly differently expressed (P_FDR_ ≤ 0.01, **STable 1**). Interestingly, most pairs of genes that showed an increase in correlation coefficient in AD had the same directional effects in gene expression; both up- or both downregulated. Vice versa, most pairs of genes that showed a decrease in correlation coefficient showed opposite directional effects in gene expression; one up- and one downregulated. Testing the association between correlation (increase/decrease) coefficient and directionality of effects (same/opposite) on gene expression resulted in a log odds ratio of 2.78 (95%CI = 2.76, 2.80, **STable 2**).

### Regulatory hubs are primarily cell type specific

Next, we constructed gene differential correlation networks for each cell type, where vertices represent genes and edges the significant differential correlations between genes (**Fig. 1b**). Degree distributions of these networks followed a power law (**SFig. 2a-g, STable 3**), showing that these networks have scale-free topology and that per network only a few central genes (hubs) are involved in the majority of altered relationships. Comparing hubs between cell types (**Fig. 1c**) showed that 824 (95%) hubs were cell type specific, and 42 (5%) hubs were shared between at least two cell types (**Fig. 3a**). Of all identified hubs (N= 866), 261 (30%) hubs had known regulatory functions; 62 (7%) were known transcription factors (TFs)^30^, 70 (8%) were known cofactors^30^ and 154 (18%) hubs were regulators of molecular functions (GO:0065009, **Fig. 3b**). Interestingly, when pairwise comparing the neighbourhoods of excitatory neuron TF hubs (N_pairs_= 930), we found 214 TF pairs with opposite differential correlations with the same genes. For example, the TF-hub *ZNF579* was negatively correlated with *CDH10* in CT (r = -0.28) and positively correlated in AD (r = 0.31, P_adj_ = 4.38 × 10^−4^). Conversely, TF-hub *ZNF33A* was positively correlated with *CDH10* in CT (r = 0.38) and negatively in AD (r = -0.23, P_adj_ = 3.29 × 10^−4^). This suggests that there are genes that are under control by different TFs in AD compared to CT. Furthermore, within the respective cell types, the neighbourhoods of 132 hubs were significantly enriched (P_FDR_ ≤ 0.01) for the KEGG AD pathway (**SFig. 3, STable 4**), namely in excitatory neurons (N= 97), inhibitory neurons (N = 33) and astrocytes (N = 2). We identified hub genes that were differentially correlated with age, including sixteen excitatory neuron hubs, two inhibitory neuron hubs (MARS and SLF2), two astrocyte hubs (ARHGEF9 and CYFIP2), and one hub from endothelial cells (SPOCK2). Additionally, we identified hub genes that were differentially correlated with Braak stages, which included one astrocyte hub (ZNF302) and three microglia hubs (ALCAM, RAB11A and RASA3).Of the cell type specific regulatory hubs, only excitatory and inhibitory neuron hubs showed functional enrichment (GO terms), albeit for distinct processes. For example, hubs of excitatory neurons were enriched for regulation of transferase activity (N = 27, P_FDR_ = 6.20 × 10^−6^, **Fig. 3c**) and negative regulation of protein phosphorylation (N = 15, P_FDR_ = 2.42 × 10^−4^), and hubs of inhibitory neurons were enriched for positive regulation of RNA biosynthetic process (N = 21, P_FDR_ = 5.65× 10^−5^, **Fig. 3d**) and regulation of transcription by RNA polymerase II (N = 24, P_FDR_ = 9.99 × 10^−5^).

**Figure 3.**
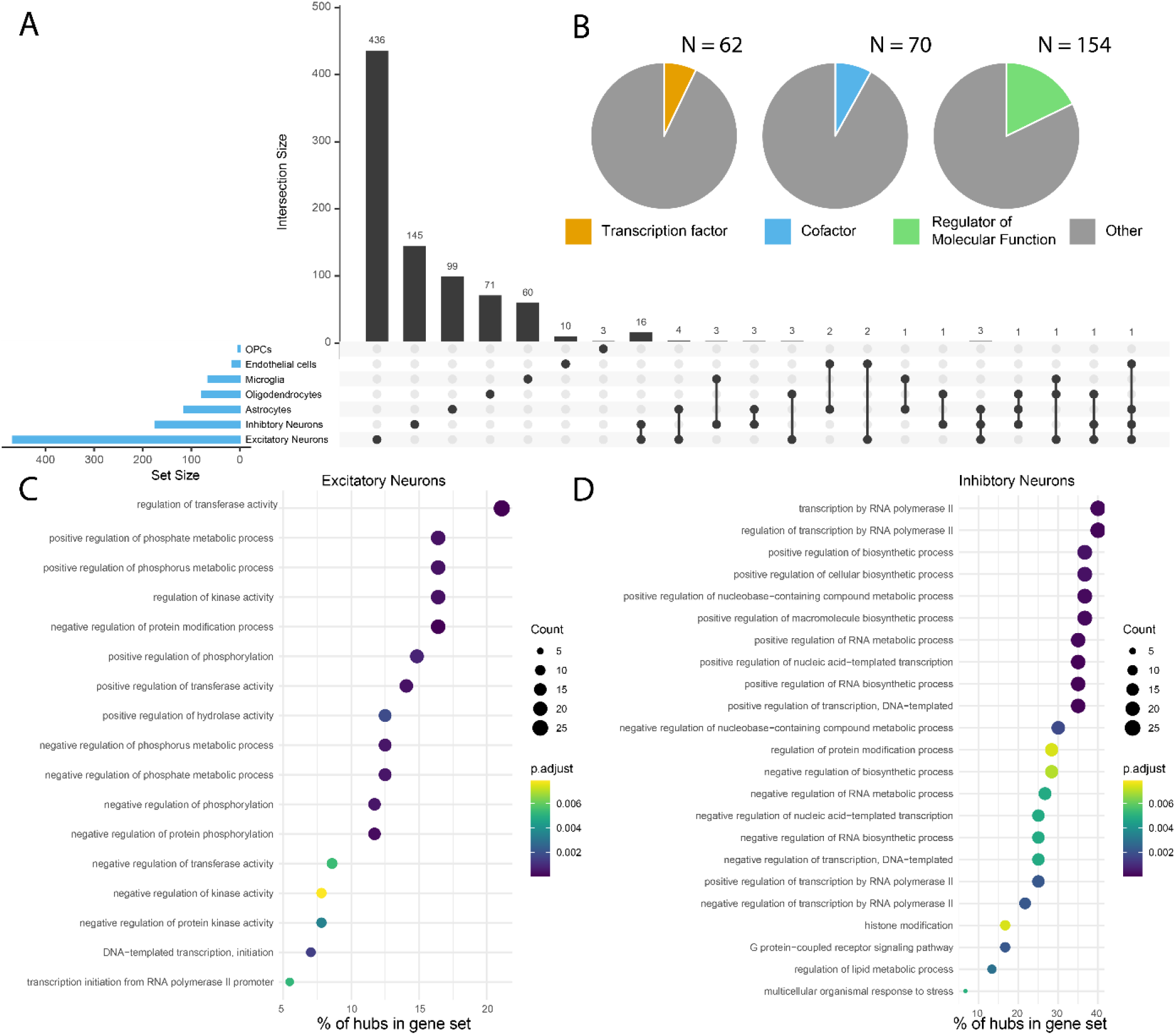
**A)** An UpSet plot of the hubs identified in each cell type, showing the degree of overlap of hubs between the cell types. **B** Pie charts showing the number of hubs that have known regulatory functions; transcription factor (orange), cofactor (blue) and regulator of molecular function (green). **C-D** Dot plot of the hub enrichment results for excitatory neurons (**C)** and inhibitory neurons (**D**), The x-axes represent the % of how many hubs belong to the GO term relative to all genes comprising the GO term. The y-axes represent the GO terms. The color of the dots represents the adjusted P-value (FDR) for the term and the size of the dots represent the number of hubs belonging respective GO term.

Next, we examined hubs that were shared between cell types (N = 42). *SARAF* was the only hub in four cell types and *DGKZ, EMC7, HNRNPH1, MOK, NDUFV3* and *S100A6* were shared hubs in three cell types. To investigate whether hubs shared between cell types also share neighbourhoods we performed a Fisher’s exact test between the neighbourhoods of the respective cell types for each hub that was shared between at least two cell types. Considering all cell types, we found 21 hubs with significantly overlapping neighbourhoods between cell types (*fisher exact test*, P_FDR_ ≤ 0.01, **SFig. 4, STable 5**). Of the 21 hubs shared between excitatory and inhibitory neurons, 19 had significantly overlapping neighbourhoods. These results show that when hub genes are shared between cell types the putative gene expression regulatory disruptions are also shared.

**Figure 4.**
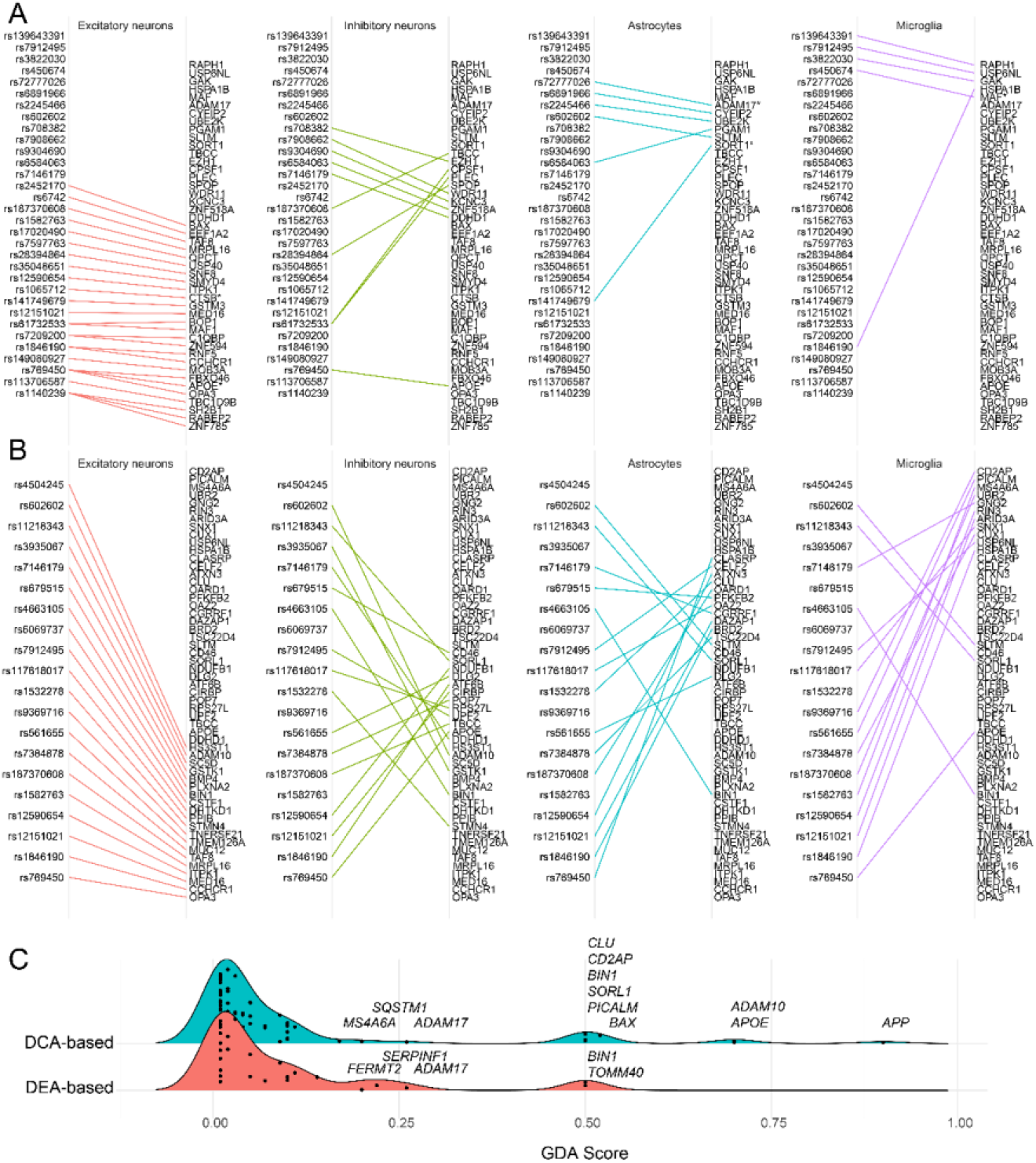
Prioritization plots of excitatory neurons, inhibitory neurons, astrocytes and microglia. **A)** hubs located nearby risk variants. **B)** The prioritized genes for the top 20 risk variants from Wightman et al (sorted on significance). **C)** Distribution of GDA scores for the DCA-based (blue) and DEA-based (red) prioritized genes.

### Differential correlation-based gene prioritization for Alzheimer’s Disease risk variants

Considering that many hubs are annotated to have known regulatory functions, we argue that the number of differential correlations can be seen as a measure of importance and could potentially hint at causal involvement of the respective hub in AD. As such, we hypothesized that the number of differential correlations can be used to prioritize genes located near AD risk variants. We focused on 79 AD risk variants identified by Wightman et al^4^ and Bellenguez et al^5^. Using CONQUER^31^, we identified 2,528 genes near these 79 variants (see methods). Of these genes, 975 were present in our collection of single cell datasets (**SFig. 5**). In total, for 32 variants at least one hub was located nearby (**Fig. 4a**, N_excitatory neurons_= 19, N_inhibitory neurons_= 9, N_astrocytes_= 6, N_oligodendrocytes_= 3, N_microglia_= 5). rs61732533 had four nearby located hubs; *MAF1* and *BOP1* in excitatory neurons and *PLEC* and *CPSF1* in inhibitory neurons. Interestingly, a dysfunction of *PLEC* in neurons is known to be associated with tau accumulation^32^. rs769450 also had four nearby located hubs; *OPA3, APOE* and *FBXO46* in excitatory neurons and *APOE* in inhibitory neurons. For five variants the annotated gene was also a hub (rs769450 - excitatory and inhibitory neurons - *APOE*, rs1065712 - excitatory neurons - *CTSB*, rs141749679 – astrocytes - *SORT1*, rs72777026 - astrocytes - *ADAM17* and rs450674 microglia - *MAF)*.

**Figure 5:**
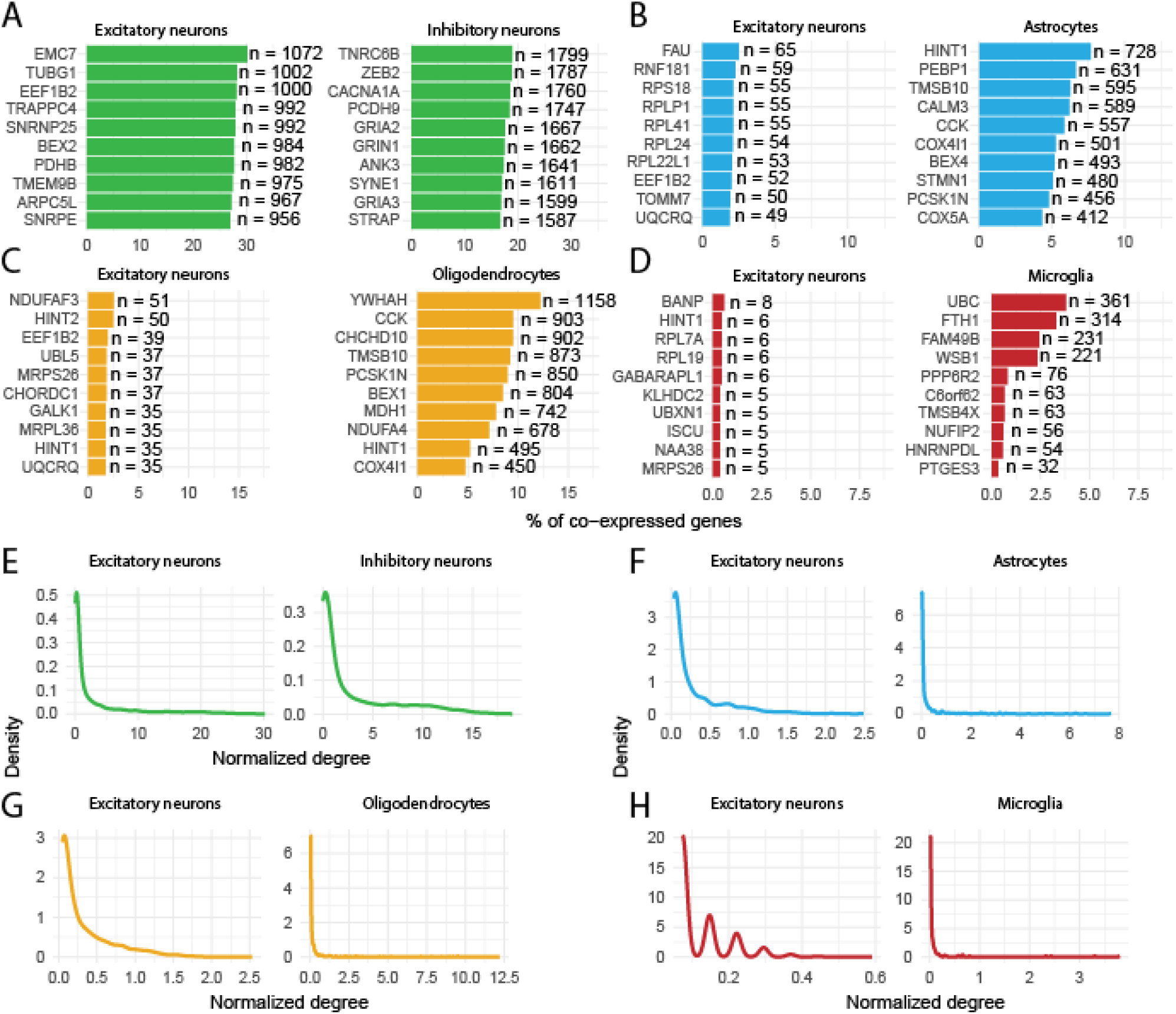
Co-expression between cell types. Top ten (defined by no. of co-expression) excitatory neuron genes co-expressed with **A)** inhibitory neuron genes, **B)** astrocyte genes, **C)** oligodendrocytes and **D)** microglia, and vice versa. E.g. EMC7 expressed in excitatory neurons is co-expressed with 1,072 genes expressed in inhibitory neurons, TNRC6B expressed in inhibitory neurons is co-expressed with 1,799 genes expressed in excitatory neurons. **E-H)** Distribution of normalized degree of **E)** genes expressed in excitatory neurons co-expressed with inhibitory neuron genes (left), and vice versa (right). **F)** Of genes expressed in excitatory neurons co-expressed with astrocyte genes, and vice versa. **G)** Of genes expressed in excitatory neurons co-expressed with oligodendrocyte genes, and vice versa. **H)** Of genes expressed in excitatory neurons co-expressed with oligodendrocyte genes, and vice versa. Degrees are normalized for total number of genes of the “other” cell type.

Next, we calculated a cell type specific normalized rank based on the number of significant differential correlations (higher number of differential correlations = higher priority). Using this rank we prioritized genes within each cell type and for each risk variant. In total, we prioritized 230 genes in all cell types (**Fig. 4b, STable 6**). For 29 variants the previously annotated gene corresponded to the highest prioritized gene in different cell types: e.g. rs769450-*APOE*, rs602602-*ADAM10*, rs4663105-*BIN1*, rs11218343-*SORL1*, rs1532278-*CLU* and *rs141749679-SORT1*. For 67 variants, another potential new risk gene ranked highest in the various cell types among which nineteen transcription factors were prioritized (e.g. rs1140239 - *ZNF785*, rs7384878 - *CUX1*). Another prioritized gene was *MAF1* for rs61732533 in excitatory neurons. *MAF1* is a stress responsive transcription factor and mTOR effector^33^. Aberrant mTOR signaling is suggested to strongly contribute to AD, mainly through oxidative stress^34^. *KCNC3* was prioritized for rs9304690 in inhibitory neurons. *KCNC3* is a potassium channel. A dysfunction of potassium channels has been associated with AD and many other neurological disorders^35^. To evaluate our prioritization approach, we compared it to prioritization using DEA. In short, using DEA we prioritized genes located nearby risk variants that were the most differentially expressed. The two sets of prioritized genes (DCA- and DEA-based) were evaluated using DisGeNET^36^. DisGeNET scores disease-gene associations (GDA, 0.01 – 0.9) based on their level of evidence in literature and from curated sources (higher GDA = more evidence). DCA- and DEA-based gene prioritization resulted in 70 and 24 genes respectively that were previously associated with AD (**Fig. 4c**). The highest scoring genes using DCA were *APP* (GDA = 0.9) and *APOE* (GDA = 0.70). The highest scoring genes using DEA were TOMM40 (GDA = 0.50) and BIN1 (GDA = 0.50). Thus, we identified more disease-associated genes and with higher levels of evidence (i.e., higher GDA score) using DCA compared to DEA, offering an internal validation that DCA can be used as a prioritization method to identify putative risk genes. Altogether, with this prioritization approach, well-known AD genes were prioritized as well as genes that have not yet been associated with AD previously. Our method hints at involvement of these lesser-known genes in AD, and as such they might be of interest for future studies.

### Co-expression analysis across cell types suggests glia-to-neuron intercellular directionality of gene expression regulation

Finally, taking advantage of the characteristics of pseudo bulk data, we explored whether co-expression of genes exceeds cell type boundaries and whether it can also be indicative of transcriptional regulation across cell types. We focused on interactions between excitatory neurons and inhibitory neurons, excitatory neurons and astrocytes, excitatory neurons and oligodendrocytes, and finally excitatory neurons and microglia. Using only CT data, co-expression between pairs of genes was assumed at a spearman rank correlation coefficient of |r| ≥ 0.6. Excitatory neurons and inhibitory neurons had the highest co-expression rate as 2.69% (n = 909,061, **Fig. 5a**) of all tested gene pairs were co-expressed. For astrocytes, oligodendrocytes and microglia this was 0.06% (n = 16,123), 0.08% (n = 16,196) and 0.02% (n= 2,078) respectively. Interestingly, when constructing co-expression graphs of each tested cell type pair we found that genes expressed in astrocytes (**Fig. 5b**), oligodendrocytes (**Fig. 5c**) and microglia (**Fig. 5d**) were more densely connected to genes in excitatory neurons, than the other way around (**Fig. 5e-h**). For example, the most densely connected gene in astrocytes, *HINT1*, was co-expressed with 728 genes in excitatory neurons, which is 7.66% of all genes measured in the excitatory neurons. The most densely connected excitatory neuron gene, *FAU (***Fig. 5b**), was co-expressed with only 65 (2.49%) genes in astrocytes. Of note, *HINT1* was only co-expressed with 238 excitatory neuron genes in the AD population, meaning it lost co-expression with 490 genes. Given that *HINT1* is implicated in Ca^2+^ signalling^37^ and that an increase of astrocytic Ca^2+^ signalling is associated with AD^38^ and thought to have downstream effects on neuronal metabolism^39^, astrocytic *HINT1* might be involved in this dysregulation. In oligodendrocytes, the most densely connected gene was *YWHAH* (n = 1,158, 12.2%), which is implicated in the regulation of many signaling pathways^40^. In microglia *UBC* (n = 361, 3.80%) was most densely connected. *UBC* is involved in ubiquitination, which is a post-translational modification process involved in the regulation of many processes^41^. The function of these densely connected genes hints at transcriptional regulation of excitatory neuron by genes expressed in inhibitory neurons, astrocytes, oligodendrocytes and microglia.

## DISCUSSION

In this study, we have provided insight into cell type specific and coordinated transcriptional changes in AD and pin-pointed putative key transcriptional regulators of the observed changes. Most importantly, we have provided a prioritization scheme that identifies probable causal genes and important cell types by superimposing the set of most differentially correlated genes onto genes located near AD risk variants. Finally, we have shown that transcriptional relationships and differences in these relationships do exceed the confines of cell types, hinting at altered intercellular communication AD. Altogether, performing differential correlation analysis (DCA) on scRNAseq data provided a comprehensive insight into transcriptional changes and consequences that are associated with AD.

Our results show that the number of altered associations a gene has with respect to other genes in the trait of interest opposed to healthy controls can be used as a measure of involvement and severity of consequence for that trait, with the respective hub as a probable key actor. In the case of AD, examples of this are *APOE, SORL1* and *ADAM10*, three well known AD genes^42,43,44,45^, which were identified as high-ranking hub genes. The ε4 allele of *APOE* is the largest contributor to genetic risk for AD^46^. Of note, *APOE* ranked especially high in neurons (both excitatory and inhibitory), while *APOE* expression is generally low in neurons. This suggest that DCA analysis of pseudo bulk data is particularly capable of also identifying novel cell type specific interactions of risk genes. Interestingly, a recent study confirmed that under stress some neurons indeed express *APOE*^47^, which might be reflected in our results.

A strength of our prioritization scheme is that it does not require expression quantitative trait loci (eQTL) or colocalization^10^ analyses for variant-gene mapping, which is generally done in GWASs. The effect of a variant on a gene is not always reflected in differential expression of the respective gene. For instance, a variant might alter the amino acid sequence of a protein, without changing the extent to which the transcript is expressed. As the function of the protein is possibly changed, it can also alter functional relationships with other proteins and their respective transcripts. Support for this is shown with the association between rs769450 (part of the ε4 allele) and *APOE*. In brain eQTL and pQTL studies^48^, rs769450 has been shown not to influence the abundance of the *APOE* protein or transcript. Additionally, in differential expression analyses of AD, *APOE* is often not among the most differentially expressed genes^18,14^. However, in our analysis, *APOE* is ranked among the most differentially corelated hub genes, highlighting the importance of looking beyond changes in expression of only one single gene at a time. Alternatively, variants might alter the enhancer or promotor regions of a gene, and as a result the respective gene might be under control of different TFs in AD compared to healthy controls. Our results indeed suggest that when comparing AD with healthy controls, there might be genes that are under transcriptional control by different TFs compared to healthy controls. Whether this is due to changes in enhancer or promotor regions remains to be elucidated.

We mainly focused on hub genes in differential correlation networks, which were defined as genes with the most (top 5%) differential correlations. As hubs are involved in the majority of altered associations, we expected these to have a regulatory function. However, a previous study evaluating differential co-expressions showed that differentially regulated targets are more likely to be identified as hubs, instead of the regulators (TFs)^28^. With this in mind, two additional layers of evidence were collected to strengthen the regulatory status of the identified hubs. First, annotations concerning a regulatory function were collected; are the hubs TFs^30^, cofactors^30^ or regulators of molecular function? The second layer of evidence was disease association; do the hubs have altered transcriptional relationships with known AD genes^49,50^? Additionally, the gene prioritization scheme provides a third layer of evidence, as it is generally assumed that the causal gene is located near the identified risk variant.

Our results showed that when a pair of genes has increased correlation in the tested group, and both genes are significantly differentially expressed, most often the genes of that pair have similar directional effects in the tested group in terms of mean expression (either up- or downregulated in both groups), whereas a decrease of the correlation coincides with opposite directional effects. However, most genes that are differentially correlated are not differentially expressed. This shows that genes that are subjected to an increase of their correlation are more likely to respond in a similar direction subjected to the perturbation, suggesting shared and altered TF control. Loss of shared control may result from regulation by other TFs. In contrast to differential expression analysis, DCA has an added value in providing a more detailed view of transcriptional changes, and whether the changes are coordinated or not.

One limitation of this study is that different brain regions were confounded by the batches, and therefore were corrected for. It is known that different brain regions have different gene co-expression networks and different cell types and cell-to-cell connectivity, and that different brain regions respond differently to AD^51^. As such, our combination of different brain regions likely favoured transcriptional changes that are shared between brain regions.

Overall, we performed DCA in single-cell data and have shown that AD is associated with coordinated transcriptional changes. Our analysis highlights the complexity and heterogeneity of cell type specific responses to AD. And lastly, with our bottom-up approach towards gene prioritization we provide a compendium of genes that could serve as guidance for functional follow-up studies.

## METHODS

### Single-cell RNAseq data

Four 10x single-cell RNAseq (scRNAseq) datasets were acquired from AMP-AD knowledge portal of which the subjects were participants of the Religious Orders Study and the Memory and Aging Project (ROS/MAP). Two 10x datasets^17,18^ were acquired from GEO (GSE157827 and GSE138852). The Seattle Alzheimer’s Disease Brain Cell Atlas (**SEA-AD**) was obtained from (https://registry.opendata.aws/allen-sea-ad-atlas/). The first dataset (***BA9***, ID: syn16780177) consisted of 24 subjects and originated from the dorsolateral prefrontal cortex (DLPFC), specifically Brodmann area 9 (BA9). Raw fastq files were obtained of this dataset. The second dataset (***BA10***, ID: syn18485175) consisted of 48 subjects and originated from the prefrontal cortex (PFC), specifically BA10. A count matrix was obtained of this dataset as it was already processed with CellRanger aligning reads to the hg38 genome^14^. The third dataset (***BA9-46***, ID: syn21670836) consisted of 32 subjects and originated from the DLPFC, BA9 and BA46. Of this dataset a count matrix was obtained as it was also processed with CellRanger aligning reads to the hg38 genome^52^. The fourth dataset (***BA9-46-Micro***, ID: syn12514624) was a microglia only dataset, consisted of 12 subjects and originated from the DLPFC, BA9 and BA46^15,16^. Of this dataset, raw fastq files were obtained. The fifth dataset (**LAU**^17^, GSE157827) consisted of 21 PFC samples and originated from 12 individuals diagnosed with AD and 9 healthy controls. Of this dataset a raw count matrix was acquired as it was also processed with CellRanger aligning reads to the hg38. The sixth dataset (**ENT**^18^, GSE138852) consisted from 12 entorhinal cortex samples and originated from 6 individuals diagnosed with AD and 6 healthy controls. Of this dataset a raw count matrix was acquired. The seventh dataset (**SEA-AD**) consisted of 89 middle temporal gyrus samples, 23 of which were diagnosed with AD and 32 were specified as CT. Of this dataset the raw count matrix was acquired.

### Clinical data

Clinical data were acquired from the AMP-AD knowledge portal (ID: syn3157322). The variable cogdx was used to characterize controls (CT), Alzheimer’s disease (AD) and other (O). Cogdx represents the clinical consensus diagnosis of cognitive status at time of death and is indicated with a value ranging from one to six. A value of one represents no cognitive impairment (CI), as such, individuals with a cogdx of one were characterized as CT. A value of four represents Alzheimer’s dementia and no other cause of CI, as such, these individuals were characterized as AD. The remaining values represent mild CI and/or other causes for dementia and these individuals were characterized as O. Besides clinical diagnosis, *APOE* genotype, Braak stage, sex, and age at time of death was also available. However, age at time of death is censored above the age 90 years. Of the **LAU, ENT** and **SEA-AD** datasets the clinical data were acquired from the corresponding sources. For both datasets; age, sex, clinical diagnosis and Braak stage were available.

### Single-cell RNA-seq data alignment and pre-processing

The two datasets (***BA9*** and ***BA9-46-Micro***) of which fastq files were acquired were processed with CellRanger (4.0.0) aligning reads to the hg38 genome, default parameters were used. Next, all datasets were jointly pre-processed. Cells with ≤20% mitochondrial counts, ≥300 total counts, ≤20,000 total counts and ≥200 measured genes, were kept for downstream analyses.

### Clustering and cell type annotation

Each dataset was separately processed for clustering and cell type annotation which was done as follows. The processed count matrix was loaded in Seurat 3.2.2^53^. The data was log-normalized, such that: 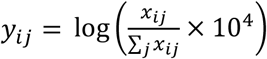,where *x*_*ij*_ and *y*_*ij*_ are the raw and normalized values for every gene *i* in every cell *j*, respectively. Next, with the 2,000 most variable genes (default with Seurat), principal components analysis (PCA) was performed. The number of principal components (PCs) used for clustering was determined using the elbow method (***BA9***:12 PCs, ***BA10***:11 PCs, ***LAU*:**11 PCs, ***BA9-46***:10 PCs, ***BA9-46-Micro:*** 7PCs, ***ENT***: 6 PCs).Next, Seurat’s `FindNeighbours`and `FindCluster`functions were used, which utilizes Louvain clustering, the resolution was set at 0.5. A UMAP plot was made to visualise and inspect the clusters. The following cell types were identified using known and previously used markers^14^: excitatory neurons (*SLC17A7, CAMK2A, NRGN*), inhibitory neurons (*GAD1, GAD2*), astrocytes (*AQP4, GFAP*), oligodendrocytes (*MBP, MOBP, PLP1*), oligodendrocyte progenitor cell (*PDGFRA, VCAN, CSPG4*), microglia (*CSF1R, CD74, C3*) and endothelial cells (*FLT1, CLDN5*). Based on differential expression of these markers between clusters, determined with Seurat’s `FindMarkers`function, cell types were assigned (SFiles. 1). When clusters were characterized by markers of multiple cell types, they were assigned as: “Unknown”. Of the **LAU** and **SEA-AD** the accompanying cell type labels were used.

### Aggregation, integration and batch correction

Per dataset, for each cell type, pseudo bulk data was generated. For instance, for each subject, cells annotated as astrocytes were aggregated in a single vector. As such, we generated cell type specific datasets. Aggregation was done based on the binary expression pattern, since the percentage of zeros for a gene in a cell population is highly associated with its mean expression^25,54^. The aggregated value of a gene for an individual was defined by the percentage of non-zero measurements in a specific cell population. Genes were kept for aggregation if they were expressed in ≥ 1% of the respective cell population in all datasets. Per cell type, the datasets were combined. Each new cell type specific dataset was batch corrected with respect to a reference dataset. First by performing a median ratio normalization^55^ and then, batch correction was performed with the `ComBat` function from the R-package sva (3.36.0)^56^. For the excitatory neurons, inhibitory neurons, astrocytes, oligodendrocytes, OPCs and endothelial cells, the ***BA9*** dataset was used as reference and for the microglia the ***BA9-46-Micro*** dataset was used as reference. Integration was confirmed with a PCA and visually inspecting the first four principal components.

### Differential correlation analysis (DCA)

Differential correlation or differential co-expression was investigated between controls and AD individuals. Differential correlation we calculated similarly as described by McKenzie et al.^29^ First, Spearman’s rank correlation coefficient between a pair of genes was calculated separately for the groups of interest based on the aggregated detection rates. This results in a correlation coefficient for each group. Next, the correlation coefficients are transformed to z-scores by means of the Fisher z-transformation^57^. Then, the difference between z-scores can be calculated with equation 1:

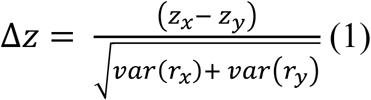

where var(r) is calculated by 1.06/(n-3), with n being the sample size of the respective groups. As Δz is normally distributed, a two-sided P-value for the differential correlation between each pair of proteins can be calculated. Besides the P-value resulting from the Z-test, empirical P-values were also calculated. The empirical null distribution was generated by permuting the group labels a 1,000 times and performing the Z-test on each pairwise combination of genes. The resulting P-values contributed to the empirical null distribution (*x*_0_). Next, the empirical P-value was calculated as:

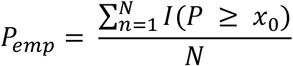

Where *I*() is an indicator function, *N* is the total number of P-values that make up the empirical null distribution and *P* is the actual P-value for which we want to determine the empirical P-value. Significance was assumed at P_emp_ ≤ 0.01.

### Classification of differential correlations

The directional change of correlation between two genes from one group to another does not reveal whether an association is lost or gained. As illustration, a change from r = -0.9 to r = -0.05 and r = 0.05 to r= 0.9 both have an increase of the correlation coefficient. However, in the first example a very strong association is lost, while in the second example a very strong association is gained. To evaluate differential correlations in terms of loss and gain of association we classified each differential correlation. First the state ***f(r)*** in both group is determined as follows:

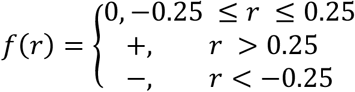

Where ***r*** is the Spearman’s rank correlation coefficient of a pair of genes in the respective group. A “**0”** represents a state of no correlation, “+**”** represents a state of positive correlation and “−**”** a state of negative correlation. When the state of a pair of genes is “−**”** or “+**”** in CT and 0 in AD, then we classify it as a *loss of association* (**-/0, +/0**). Vice versa is defined as a *gain of association* (**0/-, 0/+**) and a change from “−**”** to “+**”** or from “+**”** to “−**”**, is defined as a *flip of association* (**-/+, +/-**). Differential correlations can also remain in the same state between groups (e.g. r = 0.30 to r = 0.95), these are defined as no *change of association*.

### Differential Expression

As the aggregated detection rates were normally distributed across the individuals, differential expression analysis (DEA) was performed with a linear model, where the gene’s expression was specified as the outcome variable and the group assignment (CT = 0 and AD =1) was used as predictor variable. The function `lm` from the *stats* package from R (4.0.5) was used. Within each cell type, each gene was tested on differential expression. P-values were corrected for multiple tests, per cell type, with the Benjamini-Hochberg procedure and significance was assumed at an adjusted P-value of P_FDR_ ≤ 0.01.

### Network analysis

For each cell type, the results from the DCA were used to construct networks using igraph (1.2.6). In these networks, genes were defined as nodes and an edge between two nodes was drawn when they were significantly differentially correlated. The centrality of each gene was determined by the degree (no. of differential correlations) within the respective networks. To test whether these networks followed the power law the `fit_power_law` function from igraph was used. Hubs were defined as the top 5% of genes having the highest degree. Transcription factors and cofactors were downloaded from AnimalTFDB^30^. Genes involved in regulation of molecular functions (GO:0065009) were identified using the R-packages GOfuncR (1.10.0) and org.Hs.eg.db (3.12.0).

### GO term enrichment analysis

For each cell type, a GO term enrichment analysis was performed with the hubs that have known regulatory functions (TFs, cofactors, GO:0065009). The GO term enrichment analysis was executed with the R-package clusterProfiler (v3.18.1)^58^. Gene symbols were translated to entrez IDs making use of org.Hs.eg.db (3.12.0). For each cell type the background was defined by all genes that were present in the respective cell type specific dataset. P-values were corrected for multiple tests with the Benjamini-Hochberg procedure and significance was assumed at an adjusted P-value of P_FDR_ ≤ 0.01. The GO term regulation of molecular functions (GO:0065009) was excluded as the hubs were partly pre-filtered with this GO term.

### KEGG Alzheimer’s Disease pathway enrichment

To investigate to what degree a hub was associated with Alzheimer’s Disease, each hub was subjected to a KEGG^49^ AD pathway enrichment analysis. For each hub we performed a gene set enrichment analysis on the genes that were significantly differently correlated with the respective hub. Genes belonging to the KEGG AD pathway (ID: 05010) were defined with org.Hs.eg.db (3.12.0). Enrichment was calculated with the fisher exact test from *stats* package from R (4.0.5). P-values were corrected for multiple tests, per cell type, with the Benjamini-Hochberg procedure. For each cell type the background was defined by all genes that were present in the respective cell type specific dataset.

### Hub overlap

When hubs were identified in multiple cell types, we investigated to what degree the hubs overlap between the respective cell types, in terms of genes the hubs were differentially correlated with (e.g. are they differentially correlated with the same genes in the different cell types). This was done with the fisher exact test from *stats* package from R (4.0.5) and the background was defined by the genes that were present in both cell type specific datasets. P-values were corrected for multiple tests with the Benjamini-Hochberg procedure. A significant overlap was assumed at P-value of P_FDR_ ≤ 0.01.

### Gene prioritization

For the prioritization we started with the hubs that were identified as previously described. Next, we extracted the RS IDs from two GWASs^4,5^. In total 79 AD risk variants were extracted. For each variant, CONQUER^31^ was used to identify genes genomically located near the respective risk variants. Besides defining a fixed window of 1 Mb around the respective variant, CONQUER uses chromatin interaction to dynamically expand the search space. For each variant and cell type, genes were prioritized that were located near the respective variant and ordered based on the number of differential correlations within the respective cell type. In other words, more differential correlations, higher priority. For each of these genes, the regulatory status was evaluated (see *Network analysis*). Finally, the highest-ranking hubs were compared to the previously annotated genes, provided that the previously annotated gene was present in the respective datasets. For the variants extracted from Wightman et al^4^ the genes were assigned based on colocalization, fine-mapping and previous literature. For the variants extracted from Bellenguez et al^5^ these genes were the nearest protein coding.

## Data Availability

All data used in this study is publicly available and can be downloaded from the corresponding references.

## Availability of data and materials

The datasets used and prepared for this study can be downloaded from Zenodo (XXXX). The scripts, functions and source data for the figures are available at the Github repository: https://github.com/gbouland/xxxx.

## Author contributions

GAB, AM, and MJTR conceived the study designed the experiments. GAB performed all experiments and drafted the manuscript. GAB, KM, REK, ABS, AM, and MJTR biologically interpreted the results. GAB, KM, REK, ABS, AM, and MJTR reviewed and approved the manuscript.

## Acknowledgements

This research was supported by an NWO Gravitation project: BRAINSCAPES: A Roadmap from Neurogenetics to Neurobiology (NWO: 024.004.012). MR is recipient of ABOARD, which is a public-private partnership receiving funding from ZonMW Nationaal Dementieprogramma (#73305095007), Health∼Holland, Topsector Life Sciences & Health (PPP-allowance; #LSHM20106), as well as Edwin Bouw Fonds and Gieskes-Strijbisfonds. The results published here are in whole or in part based on data obtained from the AD Knowledge Portal (https://adknowledgeportal.org). Study data were generated from postmortem brain tissue provided by the Religious Orders Study and Rush Memory and Aging Project (ROSMAP) cohort at Rush Alzheimer’s Disease Center, Rush University Medical Center, Chicago. This work was funded by NIH grants U01AG061356 (De Jager/Bennett), RF1AG057473 (De Jager/Bennett), and U01AG046152 (De Jager/Bennett) as part of the AMP-AD consortium, as well as NIH grants R01AG066831 (Menon) and U01AG072572 (De Jager/St George-Hyslop). Study data were generated from postmortem brain tissue obtained from the University of Washington BioRepository and Integrated Neuropathology (BRaIN) laboratory and Precision Neuropathology Core, which is supported by the NIH grants for the UW Alzheimer’s Disease Research Center (P50AG005136 and P30AG066509) and the Adult Changes in Thought Study (U01AG006781 and U19AG066567). This study is supported by NIA grant U19AG060909.

## REFERENCES

1. Thies, W. & Bleiler, L. 2012 Alzheimer’s disease facts and figures. Alzheimer’s Dement. 8, 131–168 (2012).

2. Du, X., Wang, X. & Geng, M. Alzheimer’s disease hypothesis and related therapies. Transl. Neurodegener. 2018 71 7, 1–7 (2018).

3. Mohandas, E., Rajmohan, V. & Raghunath, B. Neurobiology of Alzheimer’s disease. Indian J. Psychiatry 51, 55 (2009).

4. Wightman, D. P. et al. A genome-wide association study with 1,126,563 individuals identifies new risk loci for Alzheimer’s disease. Nat. Genet. 2021 539 53, 1276–1282 (2021).

5. Bellenguez, C. et al. New insights into the genetic etiology of Alzheimer’s disease and related dementias. Nat. Genet. 2022 544 54, 412–436 (2022).

6. Holstege, H. et al. Exome sequencing identifies rare damaging variants in ATP8B4 and ABCA1 as risk factors for Alzheimer’s disease. Nat. Genet. 2022 5412 54, 1786–1794 (2022).

7. Prokopenko, D. et al. Whole-genome sequencing reveals new Alzheimer’s disease–associated rare variants in loci related to synaptic function and neuronal development. Alzheimer’s Dement. 17, 1509–1527 (2021).

8. Gallagher, M. D. & Chen-Plotkin, A. S. The Post-GWAS Era: From Association to Function. Am. J. Hum. Genet. 102, 717–730 (2018).

9. S, B. & SG, T. Interpreting findings from Mendelian randomization using the MR-Egger method. Eur. J. Epidemiol. 32, 377–389 (2017).

10. Giambartolomei, C. et al. Bayesian Test for Colocalisation between Pairs of Genetic Association Studies Using Summary Statistics. PLoS Genet. 10, e1004383 (2014).

11. Nagy, C. et al. Single-nucleus transcriptomics of the prefrontal cortex in major depressive disorder implicates oligodendrocyte precursor cells and excitatory neurons. Nat. Neurosci. 23, 771–781 (2020).

12. Wilk, A. J. et al. A single-cell atlas of the peripheral immune response in patients with severe COVID-19. Nat. Med. 26, 1070–1076 (2020).

13. Segerstolpe, Å. et al. Single-Cell Transcriptome Profiling of Human Pancreatic Islets in Health and Type 2 Diabetes. Cell Metab. 24, 593–607 (2016).

14. Mathys, H. et al. Single-cell transcriptomic analysis of Alzheimer’s disease. Nature 570, 332–337 (2019).

15. Olah, M. et al. A transcriptomic atlas of aged human microglia. Nat. Commun. 9, 1–8 (2018).

16. Olah, M. et al. Single cell RNA sequencing of human microglia uncovers a subset associated with Alzheimer’s disease. Nat. Commun. 11, 1–18 (2020).

17. Lau, S. F., Cao, H., Fu, A. K. Y. & Ip, N. Y. Single-nucleus transcriptome analysis reveals dysregulation of angiogenic endothelial cells and neuroprotective glia in Alzheimer’s disease. Proc. Natl. Acad. Sci. U. S. A. 117, 25800–25809 (2020).

18. Grubman, A. et al. A single-cell atlas of entorhinal cortex from individuals with Alzheimer’s disease reveals cell-type-specific gene expression regulation. Nat. Neurosci. 22, 2087–2097 (2019).

19. Eze, U. C., Bhaduri, A., Haeussler, M., Nowakowski, T. J. & Kriegstein, A. R. Single-cell atlas of early human brain development highlights heterogeneity of human neuroepithelial cells and early radial glia. Nat. Neurosci. 24, 584–594 (2021).

20. Squair, J. W. et al. Confronting false discoveries in single-cell differential expression. Nat. Commun. 2021 121 12, 1–15 (2021).

21. Crowell, H. L. et al. muscat detects subpopulation-specific state transitions from multi-sample multi-condition single-cell transcriptomics data. Nat. Commun. 11, 1–12 (2020).

22. Kang, H. M. et al. Multiplexed droplet single-cell RNA-sequencing using natural genetic variation. Nat. Biotechnol. 36, 89–94 (2018).

23. Van Der Wijst, M. G. P. et al. Single-cell RNA sequencing identifies celltype-specific cis-eQTLs and co-expression QTLs. Nat. Genet. 50, 493–497 (2018).

24. Pratapa, A., Jalihal, A. P., Law, J. N., Bharadwaj, A. & Murali, T. M. Benchmarking algorithms for gene regulatory network inference from single-cell transcriptomic data. Nat. Methods 2020 172 17, 147–154 (2020).

25. Svensson, V. Droplet scRNA-seq is not zero-inflated. Nature Biotechnology vol. 38 147–150 at https://doi.org/10.1038/s41587-019-0379-5 (2020).

26. Skinnider, M. A., Squair, J. W. & Foster, L. J. Evaluating measures of association for single-cell transcriptomics. Nat. Methods 16, 381–386 (2019).

27. Sanchez-Taltavull, D. et al. Bayesian correlation is a robust gene similarity measure for single-cell RNA-seq data. NAR Genomics Bioinforma. 2, (2020).

28. Bhuva, D. D., Cursons, J., Smyth, G. K. & Davis, M. J. Differential co-expression-based detection of conditional relationships in transcriptional data: Comparative analysis and application to breast cancer. Genome Biol. 20, 236 (2019).

29. McKenzie, A. T., Katsyv, I., Song, W. M., Wang, M. & Zhang, B. DGCA: A comprehensive R package for Differential Gene Correlation Analysis. BMC Syst. Biol. 10, (2016).

30. Hu, H. et al. AnimalTFDB 3.0: A comprehensive resource for annotation and prediction of animal transcription factors. Nucleic Acids Res. 47, D33–D38 (2019).

31. Bouland, G. A. et al. CONQUER: an interactive toolbox to understand functional consequences of GWAS hits. NAR Genomics Bioinforma. 2, (2020).

32. Valencia, R. G. et al. Plectin dysfunction in neurons leads to tau accumulation on microtubules affecting neuritogenesis, organelle trafficking, pain sensitivity and memory. Neuropathol. Appl. Neurobiol. 47, 73–95 (2021).

33. Cai, Y. & Wei, Y. H. Stress resistance and lifespan are increased in C. elegans but decreased in S. cerevisiae by mafr-1/maf1 deletion. Oncotarget 7, 10812 (2016).

34. Perluigi, M., Di Domenico, F., Barone, E. & Butterfield, D. A. mTOR in Alzheimer disease and its earlier stages: Links to oxidative damage in the progression of this dementing disorder. Free Radic. Biol. Med. 169, 382–396 (2021).

35. Villa, C., Suphesiz, H., Combi, R. & Akyuz, E. Potassium channels in the neuronal homeostasis and neurodegenerative pathways underlying Alzheimer’s disease: An update. Mech. Ageing Dev. 185, 111197 (2020).

36. Piñero, J., Saüch, J., Sanz, F. & Furlong, L. I. The DisGeNET cytoscape app: Exploring and visualizing disease genomics data. Comput. Struct. Biotechnol. J. 19, 2960–2967 (2021).

37. Linde, C. I., Feng, B., Wang, J. B. & Golovina, V. A. Histidine triad nucleotide-binding protein 1 (HINT1) regulates Ca2+ signaling in mouse fibroblasts and neuronal cells via store-operated Ca2+ entry pathway. Am. J. Physiol. - Cell Physiol. 304, C1098 (2013).

38. Kuchibhotla, K. V., Lattarulo, C. R., Hyman, B. T. & Bacskai, B. J. Synchronous hyperactivity and intercellular calcium waves in astrocytes in Alzheimer mice. Science (80-.). 323, 1211–1215 (2009).

39. Åbjørsbråten, K. S. et al. Impaired astrocytic Ca2+ signaling in awake-behaving Alzheimer’s disease transgenic mice. Elife 11, (2022).

40. Sato, S., Fujita, N. & Tsuruo, T. Regulation of kinase activity of 3-phosphoinositide-dependent protein kinase-1 by binding to 14-3-3. J. Biol. Chem. 277, 39360–39367 (2002).

41. Popovic, D., Vucic, D. & Dikic, I. Ubiquitination in disease pathogenesis and treatment. Nat. Med. 2014 2011 20, 1242–1253 (2014).

42. Kim, J., Basak, J. M. & Holtzman, D. M. The Role of Apolipoprotein E in Alzheimer’s Disease. Neuron vol. 63 287–303 at https://doi.org/10.1016/j.neuron.2009.06.026 (2009).

43. Kanekiyo, T., Xu, H. & Bu, G. ApoE and Aβ in Alzheimer’s disease: Accidental encounters or partners? Neuron vol. 81 740–754 at https://doi.org/10.1016/j.neuron.2014.01.045 (2014).

44. Yin, R. H., Yu, J. T. & Tan, L. The Role of SORL1 in Alzheimer’s Disease. Molecular Neurobiology vol. 51 909–918 at https://doi.org/10.1007/s12035-014-8742-5 (2015).

45. Kunkle, B. W. et al. Genetic meta-analysis of diagnosed Alzheimer’s disease identifies new risk loci and implicates Aβ, tau, immunity and lipid processing. Nat. Genet. 51, 414–430 (2019).

46. Gatz, M. et al. Role of genes and environments for explaining Alzheimer disease. Arch. Gen. Psychiatry 63, 168–174 (2006).

47. Zalocusky, K. A. et al. Neuronal ApoE upregulates MHC-I expression to drive selective neurodegeneration in Alzheimer’s disease. Nat. Neurosci. 24, 786–798 (2021).

48. Robins, C. et al. Genetic control of the human brain proteome. Am. J. Hum. Genet. 108, 400–410 (2021).

49. Kanehisa, M. & Goto, S. KEGG: Kyoto Encyclopedia of Genes and Genomes. Nucleic Acids Research vol. 28 27–30 at https://doi.org/10.1093/nar/28.1.27 (2000).

50. Kanehisa, M., Furumichi, M., Sato, Y., Ishiguro-Watanabe, M. & Tanabe, M. KEGG: Integrating viruses and cellular organisms. Nucleic Acids Res. 49, D545–D551 (2021).

51. Lancour, D. et al. Analysis of brain region-specific co-expression networks reveals clustering of established and novel genes associated with Alzheimer disease. Alzheimer’s Res. Ther. 2020 121 12, 1–11 (2020).

52. Zhou, Y. et al. Human and mouse single-nucleus transcriptomics reveal TREM2-dependent and TREM2-independent cellular responses in Alzheimer’s disease. Nat. Med. 26, 131–142 (2020).

53. Stuart, T. et al. Comprehensive Integration of Single-Cell Data. Cell 177, 1888–1902.e21 (2019).

54. Bouland, G. A., Mahfouz, A. & Reinders, M. J. T. Consequences and opportunities arising due to sparser single-cell RNA-seq datasets. Genome Biol. 2023 241 24, 1–10 (2023).

55. Anders, S. & Huber, W. Differential expression analysis for sequence count data. Genome Biol. 11, R106 (2010).

56. Leek, J. T. et al. The sva package for removing batch effects and other unwanted variation in high-throughput experiments. Bioinforma. Appl. NOTE 28, 882–883 (2012).

57. Fisher, R. A. Frequency Distribution of the Values of the Correlation Coefficient in Samples from an Indefinitely Large Population. Biometrika 10, 507 (1915).

58. Yu, G., Wang, L.-G., Han, Y. & He, Q.-Y. clusterProfiler: an R Package for Comparing Biological Themes Among Gene Clusters. Omi. A J. Integr. Biol. 16, 284–287 (2012).

