## Supplemental Figures for "Single-cell RNA sequencing data reveals rewiring of transcriptional relationships in Alzheimer’s Disease associated with risk variants"

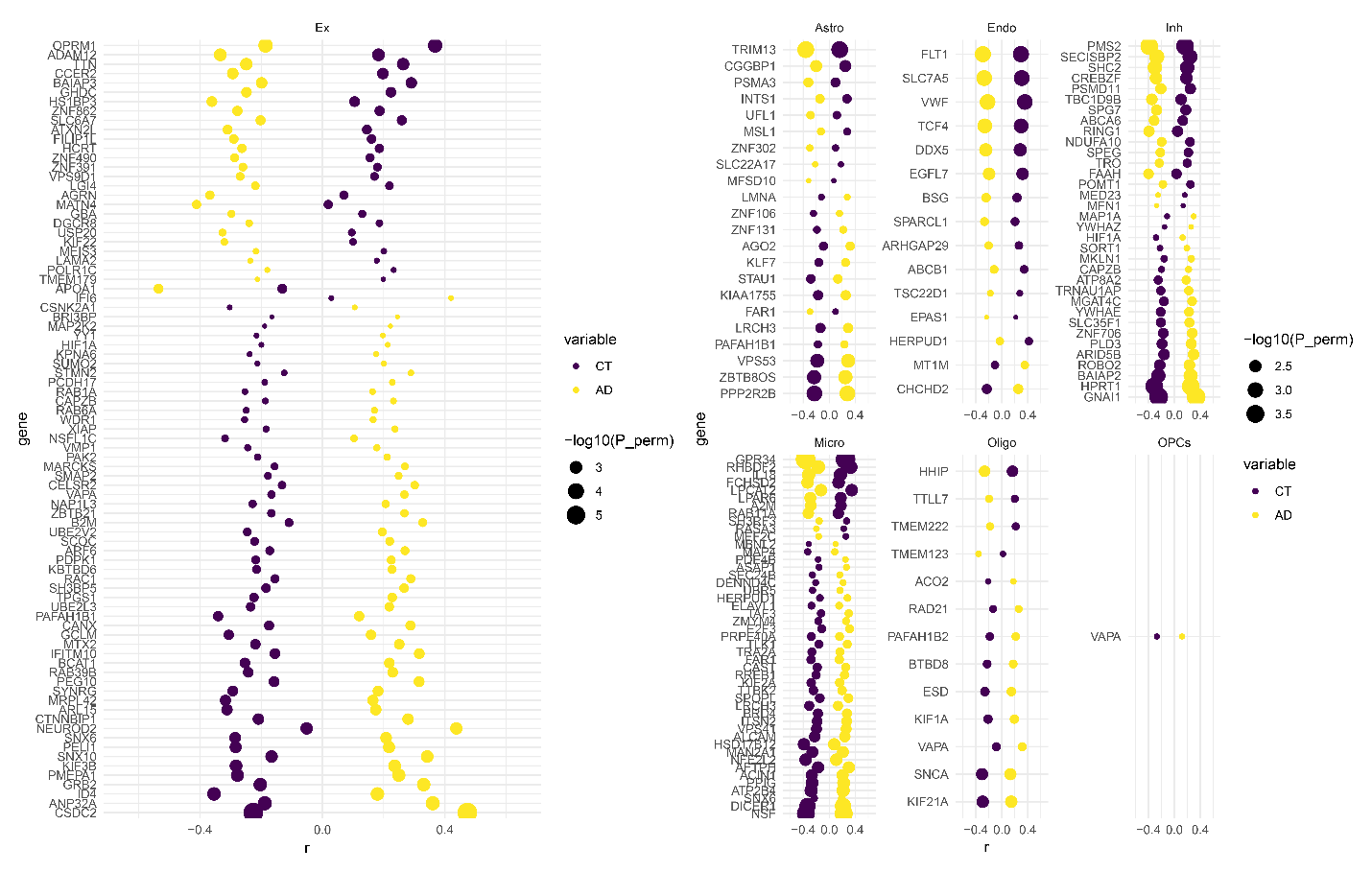
 **Supplementary Figure 1:** Differential correlation of genes with the braak stage between CT and AD individuals. X-axis represents the correlation coefficient of the gene with braak stage in the respective group (yellow = AD, purple = CT). Y-axis are the genes.

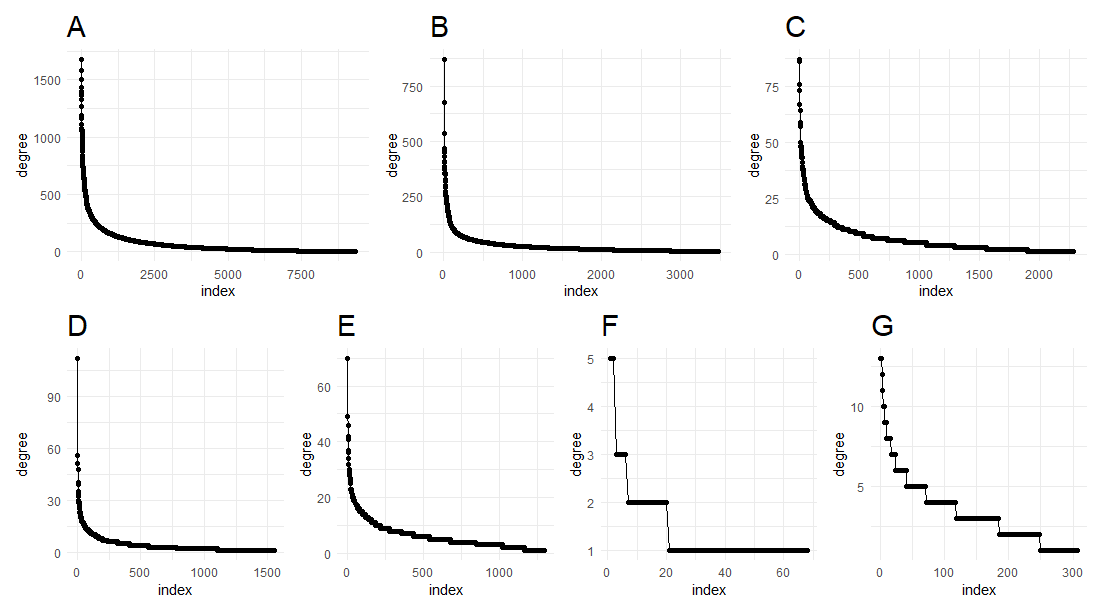

**Supplementary Figure 2:** Degree distribution of differential correlation networks of **A)** excitatory neurons, **B)** inhibitory neurons, **C)** astrocytes, **D)** oligodendrocytes, **E)** microglia, **F)** OPCs **G)** endothelial cells. Every dot is a gene, the x-axis the index of the gene sorted on degree and the y-axis is the degree.

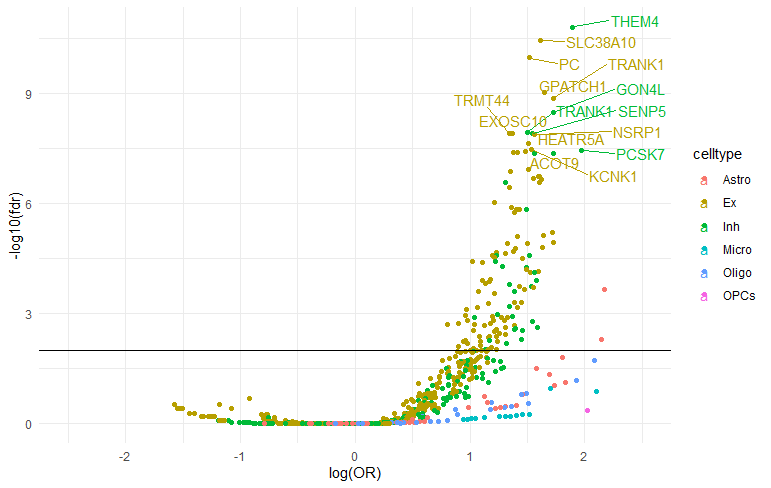
**Supplementary Figure 3:** Volcano plot of neighborhood AD enrichment. Every dot represents a hub. The x-axis represents the log odds ratio of the overlap between the hub neighborhood and the KEGG AD pathway. The y-axis represents the –log10 P_FDR_ p-value of the fisher exact test. Colors are the different cell types.

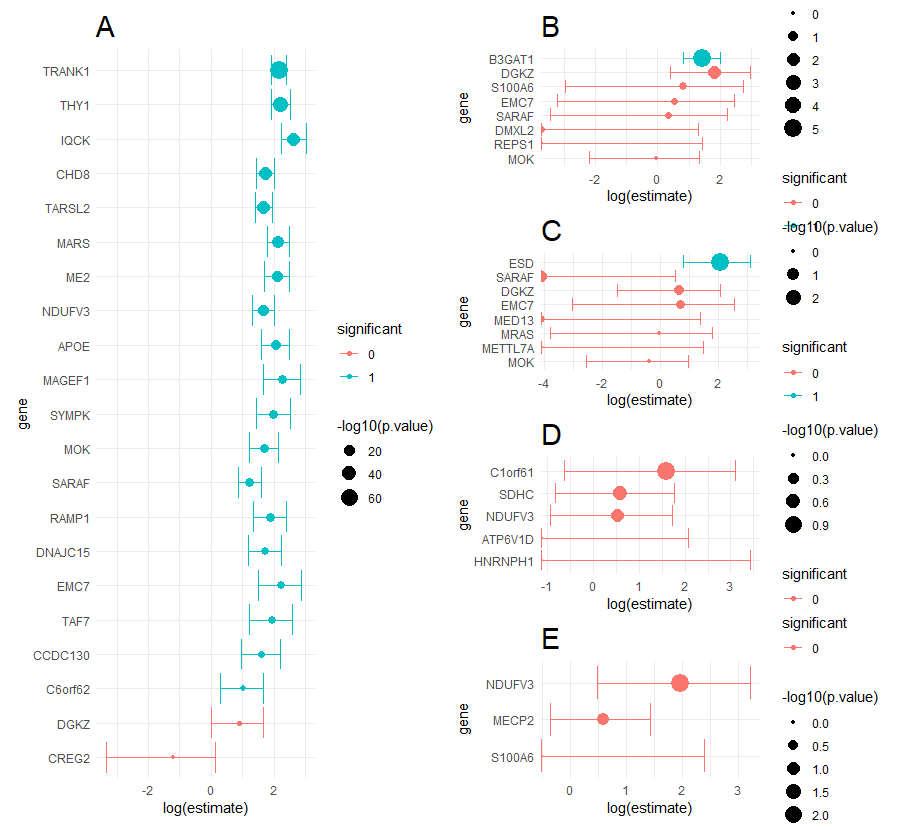

**Supplementary Figure 4:** Pairwise neighborhood enrichment of shared hubs between **A)** excitatory neurons and inhibitory neurons, **B)** inhibitory neurons and astrocytes, **C)** excitatory neurons and astrocytes, **D)** excitatory neurons and oligodendrocytes, **E)** inhibitory neurons and oligodendrocytes. X-axis is the log odds ratio of the overlap between neighborhoods of the respective hub.

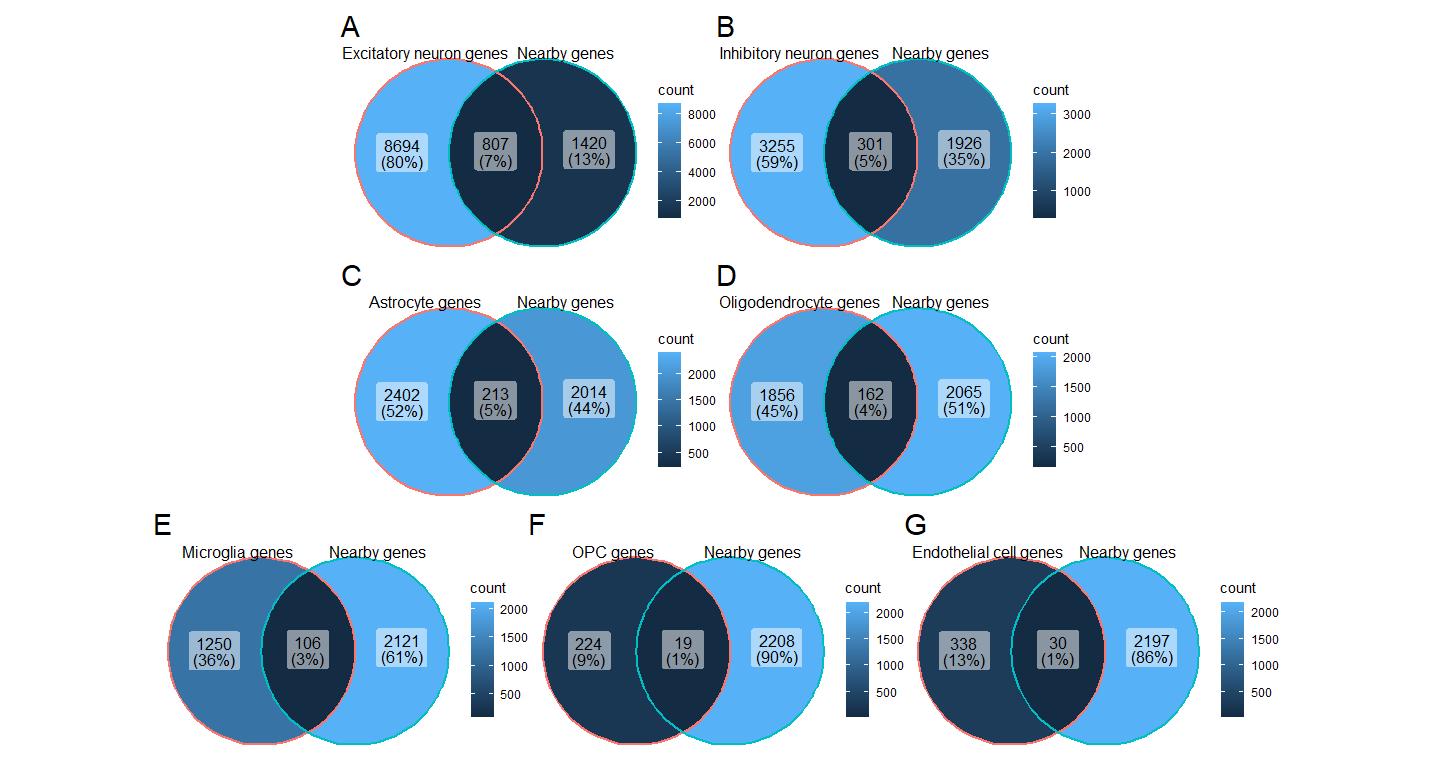

**Supplementary figure 5:** Venn diagrams of gene overlap between genes identified nearby AD risk variants and genes expressed in **A)** excitatory neurons, **B)** inhibitory neurons, **C)** astrocytes, **D)** oligodendrocytes, **E)** microglia, **F)** OPCs and **G)** endothelial cells.
